# Chemsex-associated drug use amongst men and gender-diverse people having sex with men in the UK: findings from an online community survey, 2024

**DOI:** 10.64898/2026.01.23.26344697

**Authors:** George Baldry, Giulia Habib Meriggi, Dolores Mullen, Helen Corkin, Amelia Andrews, Catherine M Lowndes, David Reid, Catherine H Mercer, John Saunders, Hamish Mohammed, Dana Ogaz

**Affiliations:** Blood Safety, Hepatitis, STI & HIV Division, UK Health Security Agency, 61 Colindale Ave, London NW9 5EQ UK; The National Institute for Health and Care Research Health Protection Research Unit in Blood Borne and Sexually Transmitted Infections at University College London in partnership with the UK Health Security Agency, London, UK; Institute for Global Health, Gower Street, London, WC1E 6BT, London, UK

**Keywords:** Chemsex, GBMSM, Drug use, Sexual health

## Abstract

**Objectives:** Chemsex is the use of select psychoactive drugs to enhance sexual experiences and has been described among gay, bisexual, and other men who have sex with men (GBMSM). We aimed to characterise sexual risk, wellbeing and health-seeking behaviours among GBMSM and gender-diverse people reporting chemsex-associated drug use.

**Methods:** We analysed data from ‘Reducing inequalities in Sexual Health’ (RiiSH), an online community survey of 2,758 UK-resident men and gender-diverse people having sex with men undertaken in November-December 2024. We compared those reporting chemsex-associated drug use with those who did not, assessing sociodemographic characteristics, well-being, sexual risk behaviours and sexual health service (SHS) engagement.

**Results:** Overall, 8% (218/2,758) reported chemsex-associated drug use in the last year. A higher proportion of participants reporting chemsex-associated drug use in the last year also reported a composite measure of sexual risk based on self-reported behaviours in the previous 3-4 months (e.g. prior bacterial STI diagnosis, ≥5 male condomless anal sex partners) compared to those who did not (85% vs 61%, p<0.001). They also more frequently reported attending a SHS in the last year (81% vs 57%, p<0.001). Those reporting chemsex-associated drug use also more frequently reported a long-term limiting mental health condition (36% vs 24%, p<0.001) and poorer personal wellbeing (e.g. reporting low life satisfaction 36% vs 20%, p<0.001).

**Conclusion:** While a minority of participants in this national, community-based sample reported chemsex-associated drug use, this group had higher sexual risk and poorer indicators of wellbeing. Many participants also attended SHS, reinforcing the key supporting role of SHS for referral pathways to harm reduction support for those experiencing problematic drug use.

## Introduction

Chemsex refers to the use of drugs before or during planned sexual activity to sustain, enhance, disinhibit or facilitate the experience; it commonly involves use of gamma-hydroxybutyrate/gamma-butyrolactone (GHB/GBL), crystal methamphetamine (‘crystal meth’), or mephedrone, and is reported by some gay, bisexual, and other men who have sex with men (GBMSM) [1]. While not all drug use associated with chemsex is inherently problematic [1, 2], chemsex is associated with increased sexual risk-taking, such as condomless anal sex (CAS) and multiple concurrent sexual partners[3], which increase the risk of sexually transmitted infection (STI) and blood-borne virus (BBV) transmission[4]. Some studies also found lower levels of HIV testing among people engaging in chemsex[3]. Beyond sexual health risks, use of these drugs, particularly when injected intravenously (‘slamming’)[5], has been associated with wider harms, including substance dependence, sexual assault or abuse, and lifestyle harms, such as loss of employment [5]. Similar patterns of chemsex-associated drug use and associated health impacts in GBMSM have also been described across Europe, North America, Asia and Australia [5-7]. Thus, ongoing understanding of the picture of chemsex and access to and use of services to support those that require help is important.

Using data from a large, online cross-sectional survey conducted in 2024, we aim to estimate the prevalence of chemsex-associated[5] drug use and describe the demographic, behavioural and wellbeing characteristics of participants using these drugs among a community sample of men and gender-diverse people having sex with men in the UK.

## Methods

The ‘Reducing inequalities in Sexual Health’ (RiiSH) survey is a series of annual online cross-sectional surveys, assessing the sexual health and well-being of a UK community sample of men and gender-diverse people who have sex with men. For RiiSH-2024, participants were recruited through advertisements on social networking sites and geospatial dating platforms from 18^th^ November-11^th^ December 2024. Participants eligible to take part included self-identifying men (cisgender or transgender), transgender women or gender-diverse people who were assigned male at birth (AMAB), aged ≥16 years, residing in the UK and reporting sex with a man or non-binary AMAB person in the last year.

We examined chemsex-associated drug use in all RiiSH-2024 participants. Participants were asked if they had ever taken crystal meth, mephedrone or GHB/GBL and the recency of last use. Those who reported use in the previous 3-4 months (i.e. since August 2024) were asked what proportion of their sex involved chemsex-associated drug use on a scale from none to all. All participants were asked separately if they had ever injected drugs, excluding prescribed medicines or anabolic steroids.

We present the proportion of participants who reported chemsex-associated drug use, ever and in the last year. Differences in the sociodemographic characteristics (age-group, sexual orientation, ethnicity, area of residence, financial comfort, country of birth, education, employment), clinical history (HIV status, ever use of online postal self-sampling kits for STI testing, SHS attendance in last year), sexual risk behaviour (see below), and personal-wellbeing (life satisfaction, long-term limiting physical or mental health conditions) among those reporting and not reporting chemsex-associated drug use were examined using Pearson’s chi-squared test. Given the descriptive aim of the study and a priori concerns regarding small subgroup sizes among those reporting chemsex-associated drug use, regression modelling was considered outside the scope of the analysis.

A composite measure of sexual risk was used in analyses, defined by reporting any of the following within the last 3-4 months: a bacterial STI diagnosis (chlamydia/gonorrhoea/syphilis), ≥5 male condomless anal sex (CAS) partners, and meeting partners through sex-on-premises venues, public sex environments (PSE i.e. cruising environments), or at private sex parties, and/or HIV pre-exposure prophylaxis (PrEP) use in the last year. We also report sexual risk behaviours included in the composite measure separately.

To assess wellbeing, we used UK’s Office for National Statistics’ (ONS) personal well-being indicators, including a dichotomised measure of low life-satisfaction based on ONS harmonisation standards (i.e. low vs medium, high, very high)[8]. For participants reporting a long-term physical or mental health conditions, we derived a binary variable indicating limitations. Participants who responded ‘Yes, a little’, ‘Yes, a lot’ (vs ‘Not at all’) to the question, “Does your condition or illness reduce your ability to carry out day-to-day activities?” were classified as having a limiting condition. Those not reporting a long-term health condition or who did not indicate limitations were classified as having no limitation. We created a binary variable to assess the frequency of sex under the influence of alcohol in the last year (over half of sex vs less than half).

## Results

There were 2,758 participants included in analyses, with a median age of 45 (IQR: 36-55). Most were cisgender men (95%), gay (80%), of white ethnicity (88%), and lived in England (87%). The majority were employed (78%), and degree-educated (59%) (Table 1).

**Table 1.** Sociodemographic characteristics, sexual risk behaviours and service use in participants reporting and not reporting chemsex-associated drug use in the last year.

|  | All Participants | No chemsex-associated drug use in last year | Chemsex-associated drug use in last year | p-value |
| --- | --- | --- | --- | --- |
| Number of respondents | 2,758 (100.0%) | 2,540 (92.1%) | 218 (7.9%) |  |
| Age (years) at survey completion (median, IQR) | 45 (36-55) | 45 (36-55) | 45 (37-55) |  |
| **Sociodemographic characteristics** |  |  |  |  |
| Age-group (years) |  |  |  | 0.713 |
| 16-29 | 337 (12.2%) | 314 (12.4%) | 23 (10.6%) |  |
| 30-44 | 1,010 (36.6%) | 927 (36.5%) | 83 (38.1%) |  |
| 45 and over | 1,411 (51.2%) | 1,299 (51.1%) | 112 (51.4%) |  |
| Sexual orientation |  |  |  | 0.006 |
| Gay/homosexual | 2,207 (80.0%) | 2,017 (79.4%) | 190 (87.2%) |  |
| Bisexual, straight, or other‡ | 551 (20.0%) | 523 (20.6%) | 28 (12.8%) |  |
| Ethnic group |  |  |  | 0.063 |
| White | 2,435 (88.3%) | 2,251 (88.6%) | 184 (84.4%) |  |
| Another ethnic group | 323 (11.7%) | 289 (11.4%) | 34 (15.6%) |  |
| Area of residence |  |  |  | <0.001 |
| Outside London/unknown (England) | 1,647 (59.9%) | 1,556 (61.5%) | 91 (41.9%) |  |
| Lives in London | 748 (27.2%) | 646 (25.5%) | 102 (47.0%) |  |
| Outside England | 354 (12.9%) | 330 (13.0%) | 24 (11.1%) |  |
| Comfortable financial situation |  |  |  | <0.001 |
| No | 1,512 (54.8%) | 1,363 (53.7%) | 149 (68.3%) |  |
| Yes (top two quartiles) § | 1,246 (45.2%) | 1,177 (46.3%) | 69 (31.7%) |  |
| UK born |  |  |  | <0.001 |
| No | 579 (21.0%) | 513 (20.2%) | 66 (30.3%) |  |
| Yes | 2,179 (79.0%) | 2,027 (79.8%) | 152 (69.7%) |  |
| Degree-level education |  |  |  | 0.294 |
| No | 1,130 (41.0%) | 1,048 (41.3%) | 82 (37.6%) |  |
| Yes | 1,628 (59.0%) | 1,492 (58.7%) | 136 (62.4%) |  |
| Employed |  |  |  | 0.004 |
| No | 619 (22.4%) | 553 (21.8%) | 66 (30.3%) |  |
| Yes | 2,139 (77.6%) | 1,987 (78.2%) | 152 (69.7%) |  |
| **Clinical history** |  |  |  |  |
| HIV status |  |  |  | <0.001 |
| HIV negative/unknown | 2,458 (89.1%) | 2,298 (90.5%) | 160 (73.4%) |  |
| PLWHIV (tested positive) | 300 (10.9%) | 242 (9.5%) | 58 (26.6%) |  |
| Ever used online self-sampling kits for STI testing |  |  |  | 0.192 |
| No | 1,720 (62.4%) | 1,593 (62.7%) | 127 (58.3%) |  |
| Yes | 1,038 (37.6%) | 947 (37.3%) | 91 (41.7%) |  |
| Visited SHS in last year |  |  |  | <0.001 |
| No | 1,134 (41.1%) | 1,093 (43.0%) | 41 (18.8%) |  |
| Yes | 1,624 (58.9%) | 1,447 (57.0%) | 177 (81.2%) |  |
| **Behavioural characteristics** |  |  |  |  |
| Composite measure of sexual risk ¶ |  |  |  | <0.001 |
| No | 1,035 (37.5%) | 1,002 (39.4%) | 33 (15.1%) |  |
| Yes | 1,723 (62.5%) | 1,538 (60.6%) | 185 (84.9%) |  |
| PrEP use in the last year † |  |  |  | <0.001 |
| No PrEP use in last year | 1,215 (49.4%) | 1,179 (51.3%) | 36 (22.5%) |  |
| PrEP use in last year | 1,243 (50.6%) | 1,119 (48.7%) | 124 (77.5%) |  |
| Number of male condomless anal sex partners (CAS) in last 3-4 months |  |  |  | <0.001 |
| <5 CAS partners | 2,086 (75.6%) | 1,981 (78.0%) | 105 (48.2%) |  |
| ≥5 CAS partners | 672 (24.4%) | 559 (22.0%) | 113 (51.8%) |  |
| Bacterial STI diagnosis in last 3-4 months ‡‡ |  |  |  | <0.001 |
| No | 2,487 (90.2%) | 2,329 (91.7%) | 158 (72.5%) |  |
| Yes | 271 (9.8%) | 211 (8.3%) | 60 (27.5%) |  |
| Met partners at sex-on-premises venues, cruising sites or sex parties in last 3-4 months |  |  |  | <0.001 |
| No | 1,847 (67.0%) | 1,746 (68.7%) | 101 (46.3%) |  |
| Yes | 911 (33.0%) | 794 (31.3%) | 117 (53.7%) |  |
| Ever injected any drug other than prescribed medicines or anabolic steroids |  |  |  | <0.001 |
| No | 2,648 (96.0%) | 2,500 (98.4%) | 148 (67.9%) |  |
| Yes | 110 (4.0%) | 40 (1.6%) | 70 (32.1%) |  |
| Frequency of sex under the influence of alcohol in the last year |  |  |  | 0.004 |
| Less than half or none | 2,443 (88.6%) | 2,263 (89.1%) | 180 (82.6%) |  |
| At least half | 315 (11.4%) | 277 (10.9%) | 38 (17.4%) |  |
| **Wellbeing** |  |  |  |  |
| Long-term limiting mental health condition |  |  |  | <0.001 |
| No | 2,078 (75.3%) | 1,939 (76.3%) (79.2%) | 139 (63.8%) |  |
| Yes | 680 (24.7%) | 601 (23.7%) | 79 (36.2%) |  |
| Long-term limiting physical health condition |  |  |  | 0.167 |
| No | 2,176 (78.9%) | 2,012 (79.2%) | 164 (75.2%) |  |
| Yes | 582 (21.1%) | 528 (20.8%) | 54 (24.8%) |  |
| Low life satisfaction |  |  |  | <0.001 |
| No (medium, high, very high satisfaction) | 2,159 (78.4%) | 2,019 (79.6%) | 140 (64.2%) |  |
| Yes | 594 (21.6%) | 516 (20.4%) | 78 (35.8%) |  |
| ‡Includes those identifying as bisexual, straight/heterosexual, or another way. §Top two quartiles ("I am comfortable"/"I am very comfortable" from the question, "How would you best describe your current financial situation". ¶ Composite measure of sexual risk defined by reporting any of the following within the last 3-4 months: a bacterial STI diagnosis (chlamydia/gonorrhoea/syphilis), having ≥5 condomless anal (CAS) sex partners, and meeting partners through sex-on-premises venues, public sex environments (i.e. cruising environments), or at private sex parties, and/or HIV pre-exposure prophylaxis (PrEP) use in the last year. † Reported in those no known to be HIV positive. ‡‡ Diagnosis of chlamydia, gonorrhoea, and/or syphilis. PrEP=HIV pre-exposure prophylaxis. PLWHIV=people living with HIV. STI=sexually transmitted infection. SHS=sexual health service. CAS=condomless anal sex. RiSH='Reducing inequalities in Sexual Health'. Chemsex-associated drug use=self-reported of crystal methamphetamine, mephedrone or gamma-hydroxybutyrate/gamma-butyrolactone. |  |  |  |  |

Overall, 15% (408/2,758) reported chemsex-associated drug use ever in their lifetime, and 8% (218/2,758) reported use in the last year (or 53% [218/408] of those reporting use ever). Among 138 participants reporting chemsex-associated drug use in the last 3-4 months, 90% (123/136, where specified) indicated that they used these drugs during some of their sexual encounters over the same period; among whom, 65% (80/123) reported use in at least half of the sexual encounters they had. Among all participants, 4% (110/2,758) reported ever injecting any non-prescribed drugs, with 41% of these participants (45/110) reporting that they had injected drugs in the in the last 3-4 months.

The median age of those reporting chemsex-associated drug use in the last year (n=218) was 45 years (interquartile range [IQR]: 37-54), similar to those not reporting use (45 [IQR:36-55]) (Table 1). A higher proportion of those reporting chemsex-associated drug use identified as gay (87% vs 80% in those reporting no use, p=0.006), resided in London (47% vs 26%, p<0.001), and were living with HIV (27% vs 10%, p<0.001). A smaller proportion also reported being financially comfortable (32% vs 46%, p<0.001), and people using chemsex-associated drugs also more commonly reported having a limiting long-term mental health condition (36% vs 24%, p<0.001) or low life satisfaction (36% vs 20%, p<0.001).

Participants reporting chemsex-associated drug use in the last year more frequently reported our composite measure of sexual risk (85% vs 61%, p<0.001) and attending a SHS in the last year (81% vs 57%, p<0.001). Chemsex-associated drug use was also more frequently reported in those reporting PrEP use in the last year (78% vs 49%, p<0.001, in those not known to be HIV positive). Reporting that at least half of their sexual encounters in the last year was under the influence of alcohol was also more common in this group (17% vs 11%, p=0.004).

## Discussion

While fewer than 1 in 10 men and gender-diverse people having sex with men reported chemsex-associated drug use in the previous year, this group had high levels of sexual risk and poorer wellbeing, including limiting long-term mental health conditions. Findings affirm that chemsex-associated drug use remains an important public health issue given a range of intersectional vulnerabilities and need for tailored harm reduction and holistic support.

The prevalence of chemsex-associated drug use (15% ever, 8% in the last year) and co-occurring characteristics appear consistent with reporting prior to the widespread disruptions caused by COVID-19 in similar community samples [3, 9]. Chemsex-associated drug use was more common among participants who identified as gay, those residing in London, and among those living with HIV. Nearly half of participants reporting use lived in London, aligning with prior London-focused studies [10, 11] and providing ongoing evidence of higher prevalence in urban settings [12, 13]. Patterns of use may reflect the influence of higher numbers of GBMSM residing in larger UK cities [14], and established sexual networks that use chemsex-associated drugs. Although our survey collected information on participants’ area of residence, we were unable to distinguish rural and urban settings. Consequently, we were unable to understand how area of residence or location of sexual activity was associated with chemsex-associated drug use.

These findings demonstrate a range of potential negative outcomes that may accompany chemsex-associated drug use, alongside patterns of risk-taking behaviours and potential wellbeing needs. Participants who reported chemsex-associated drug use also reported higher levels of sexual risk behaviours, including condomless anal sex partnerships, and meeting partners at sex-on-premise venues[2]). This group also reported lower wellbeing indicators, such as low life satisfaction, and having a limiting long-term mental health condition. A higher frequency of sex under the influence of alcohol was also more common in those reporting chemsex-associated drug use. These factors may act either as drivers or consequences of concurrent sex and drug use[2]. Irrespective of directionality, they could indicate persistent intersectional vulnerabilities, which are important factors to consider when designing and implementing effective interventions for those reporting chemsex-associated drug use.

Those reporting chemsex-associated drug use had greater engagement with SHS in the last year and higher levels of PrEP use. Similar self-mitigation of risk is consistently reported across literature [2, 3, 15], including higher levels of STI testing, PrEP use, and intentional structuring of drug use (e.g. dosage control and avoidance of injecting practices). While these findings suggest that those using chemsex drugs are aware of their elevated risk and seek to mitigate it through SHS engagement, their service use may still be driven by their sexual health needs, not directly related to chemsex-associated drug use.

### Implications of findings

High engagement with SHS among participants reporting chemsex-associated drug use suggests the importance of these settings as key points of contact for assessing and addressing chemsex-related care needs. Qualitative work in England indicates that SHS are often preferred by service users over traditional drug misuse services, which may be perceived as more stigmatising[16]. Greater shifts towards digital sexual health service delivery may further complicate appropriate support or referral for chemsex-related needs, and should consider appropriate triage[17].

Although chemsex-associated drug use was more commonly reported in London, our findings indicate chemsex-associated drug use across the UK. It is therefore essential to also ensure adequate healthcare and social infrastructure to meet the needs of people using chemsex-associated drugs throughout the UK which minimises individual and policy-level barriers. Holistic, culturally-competent care, including mental health and psychosocial interventions, that address stigma and shame associated with seeking help for problematic chemsex-associated drug use could play a role in meeting the needs of this population. Support approaches could include harm reduction education e.g. safer injection practices and safer use of substances such as GHB, while fostering sex positivity that promotes intimacy, connection and pleasure [15].

### Strengths and Limitations

We used data from a convenience sample of people recruited using social media and dating applications, where participants are more likely to report sexual risk behaviours than the general GBMSM population[18]. Our estimate of chemsex-associated drug use (ever) may be overestimated given the older age-distribution of participants. However, drawing from a community sample may enhance generalisability compared to studies relying solely on SHS recruitment. Given the small number of participants reporting chemsex-associated drug use, adjusted analyses to account for confounding were not performed. The RiiSH survey did not collect any identifiers such as name or address to reduce social desirability bias, and this may have facilitated reporting of reported chemsex-associated drug use and injection practices. Still, this study lacks diversity in key demographic areas (e.g. ethnicity, age, sexual identity) limiting our ability to assess chemsex-associated drug use in more marginalised groups.

## Conclusion

While the proportion of men and gender-diverse people engaging in chemsex-associated drug use was relatively low, the more frequently reported levels of sexual risk behaviours, which can increase STI and BBV transmission risk, highlights the need for targeted interventions. Patterns of use and reported risks appear largely consistent with studies conducted before the COVID-19 pandemic. Effective harm reduction must tackle a wide spectrum of needs, including general and sexual wellbeing, and mental health. These efforts must be implemented within diverse healthcare settings and not be lost in digital delivery.

Given the intricate sociocultural landscape in which chemsex occurs, interventions to reduce the public health impact of associated harms should be built collaboratively, incorporating meaningful community coproduction. This can empower people to enhance sexual wellbeing, reduce harm, and access appropriate care, while also strengthening the capacity of SHS and voluntary sector organisations to respond to needs appropriately within the UK and internationally.

## Acknowledgements

The authors wish to thank all participants to RiiSH 2024 and Takudzwa Mukiwa (Terrence Higgins Trust) for contributions to survey implementation. Authors acknowledge the members of the National Institute for Health and Care Research Health Protection Research Unit (NIHR HPRU) in Blood Borne and Sexually Transmitted Infections (BBSTI) Steering Committee: Professor Caroline Sabin (HPRU Director), Dr John Saunders (UKHSA Lead), Professor Catherine H Mercer, Professor Gwenda Hughes, Dr Hamish Mohammed, Professor Greta Rait, Dr Ruth Simmons, Professor William Rosenberg, Dr Tamyo Mbisa, Professor Rosalind Raine, Dr Sema Mandal, Dr Rosamund Yu, Dr Samreen Ijaz, Dr Fabiana Lorencatto, Dr Rachel Hunter, Dr Kirsty Foster and Dr Mamoona Tahir.

## Ethical considerations

Ethical approval of this study was provided by the UKHSA Research and Ethics Governance Group (REGG; ref: R&D 524).

## Consent to participate

Online informed consent was received from all participants and all methods were performed in accordance with guidelines and regulations set by the UKHSA REGG.

## Data availability

The data that support the findings of this study are not publicly available to protect participant privacy. However, some aggregate data are available upon reasonable request from the UK Health Security Agency (UKHSA). Requests can be directed to.

## Contributions

DM, GB, CML, DR, CHM, JS, HM, DO reviewed and updated the survey instrument. Survey implementation, data collection and data management were carried out by DM, GB, DO. GB, GHM, HM, DO curated secondary analysis plan with review and contributions from all authors. GB and GHM conducted analysis and completed the first manuscript draft. All authors contributed to successive drafts and reviewed and approved the final manuscript.

